# Electrocardiogram and Photoplethysmogram-based Heart Rate Variability Are Not Equivalent: A Bayesian Simulation Analysis

**DOI:** 10.1101/2023.08.24.23294449

**Authors:** Hayden G Dewig, Jeremy N Cohen, Eric J Renaghan, Miriam E Leary, Brian K Leary, Jason S. Au, Matthew S Tenan

**Affiliations:** Rockefeller Neuroscience Institute, West Virginia University, Morgantown, WV; Department of Kinesiology and Health Sciences, University of Waterloo, Waterloo, ON, Canada; Department of Athletics, University of Miami, Miami, FL; Division of Exercise Physiology, West Virginia University, WV

## Abstract

**Background:** Heart rate variability (HRV) is a common measure of autonomic and cardiovascular system function assessed via electrocardiography (ECG). Consumer wearables, commonly employed in epidemiological research, use photoplethysmography (PPG) to report HRV metrics (PRV), although these may not be equivalent. One potential cause of dissociation between HRV and PRV is the variability in pulse transit time (PTT). This study sought to determine if PPG-derived HRV (i.e., PRV) is equivalent to ECG-derived HRV and ascertain if PRV measurement error is sufficient for a biomarker separate from HRV.

**Methods:** The ECG data from 1,084 subjects were obtained from the PhysioNet Autonomic Aging dataset, and individual PTT variances for both the wrist (n=42) and finger (n=49) were derived from Mol et al. A Bayesian simulation was constructed whereby the individual arrival times of the PPG wave were calculated by placing a Gaussian prior on the individual QRS-wave timings of each ECG series. The standard deviation of the prior corresponds to the PTT variances. This was simulated 10,000 times for each PTT variance. The root mean square of successive differences (RMSSD) and standard deviation of N-N intervals (SDNN) were calculated for both HRV and PRV. The Region of Practical Equivalence bounds (ROPE) were set a priori at ±0.2% of true HRV. The Highest Density Interval (HDI) width, encompassing 95% of the posterior distribution, was calculated for each PTT variance.

**Results:** The lowest PTT variance (2.0 SD) corresponded to 88.4% within ROPE for SDNN and 21.4% for RMSSD. As the SD of PTT increases, the equivalence of PRV and HRV decreases for both SDNN and RMSSD. Thus, between PRV and HRV, RMSSD is nearly never equivalent and SDNN is only somewhat equivalent under very strict circumstances. The HDI interval width increases with increasing PTT variance, with the HDI width increasing at a higher rate for RMSSD than SDNN.

**Conclusions:** For individuals with greater PTT variability, PRV is not a surrogate for HRV. When considering PRV as a unique biometric measure, our findings reveal that SDNN has more favorable measurement properties than RMSSD, though both exhibit a non-uniform measurement error.

## Introduction

Heart rate variability (HRV) is commonly used as an index of autonomic function (1, 2) and is often extrapolated to whole-body health or all-cause mortality risk (3–5). HRV is classically defined by beat-to-beat fluctuations in the time between QRS complexes (i.e., ventricular excitation) in the electrocardiogram (ECG), also called the R-R interval (6). From the R-R interval, both time- and frequency-domain variables can be derived, and researchers often ascribe characteristics to these measures such as sympathetic function, parasympathetic function or vagal tone (2, 6, 7). Most commonly, HRV has been used as a biomarker to determine cardiovascular disease risks, responses to stress, and training modulations (4, 8–11).

HRV has become an attractive measure within the health and fitness industries because of its claims to monitor training adaptations and “physiological readiness.” Many wearable health monitoring devices (e.g., watches, rings) advertise their capability to measure “heart rate variability.” These devices, in nature, are located peripherally and most commonly use photoplethysmography (PPG) to determine pulse rate intervals at the finger or wrist (12–14). PPG assesses changes in blood volume in the peripheral tissues via infrared light, which is a functional product of cardiac electrical activity after a short mechanical delay, from cardiac excitation to blood ejection to arrival at the distal tissue. Therefore, the outcome of this PPG detection method is more appropriately termed pulse rate variability (PRV), rather than HRV (15, 16). Although the calculations for the variability indices are similar (e.g., root mean square of successive differences, standard deviation of N-N intervals), the outcomes of PRV (i.e., mechanical outcome) may not truly reflect HRV (i.e., electrical outcome).

Previous studies have suggested that a number of factors can influence pulse rate independent of heart rate, such as postural, physiological, environmental, and technical considerations (17–19). For example, Yuda and colleagues (16) outlined the steps between the electrical impulse within the heart to the mechanical pulse at peripheral site, suggesting that there is room for error between the two signals. More specifically, the authors, as well as others (1, 20, 21), have indicated that pulse transit time (PTT) is one measurable factor that can, in part, explain the differences between HRV and PRV. The PTT, defined as the time from the electrical R wave to the foot of the PPG-derived pulse wave (see Figure 1), can vary beat-by-beat due to autonomic, respiratory, or other modulations (e.g., vascular branching, aging), introducing meaningful heterogeneity to the HRV-PRV relationship.

**Figure 1.**
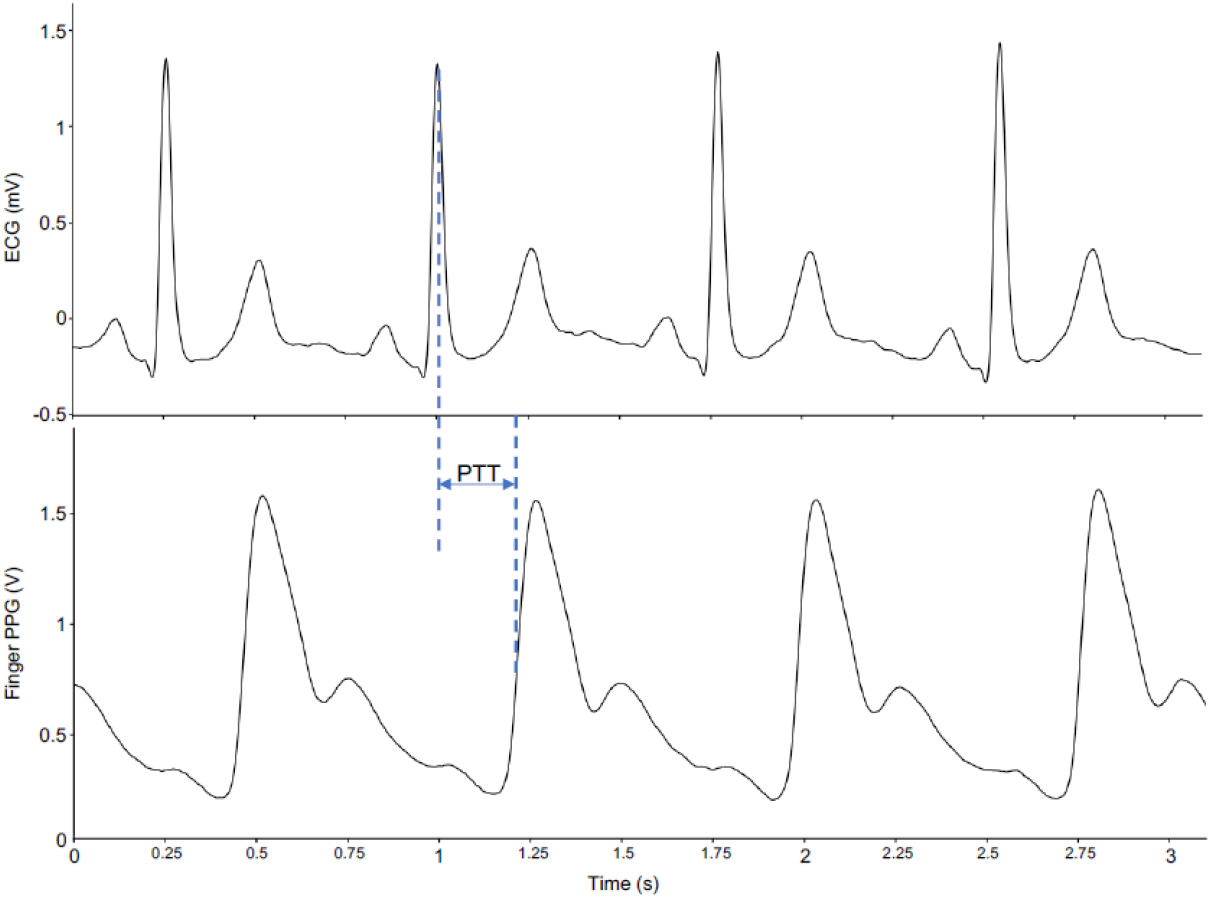
A representative example depicting the R-R intervals from an electrocardiogram (ECG) and the P-P intervals from finger photoplethysmography (PPG). Pulse transit time (PTT) is demonstrated as the time difference between the R peak and first derivative peak of the PPG signal.

Due to the influence of PTT on PRV, the relationship between ECG-derived HRV and PPG-derived PRV has been critically examined with inconsistent results. For example, Selvaraj et al. (1) found a high correlation (*r* = 0.998) between the R-R interval and P-P interval at the finger and minimal error (~0.1 ms) between variability measures (e.g., mean NN interval). Further, Gil and colleagues (21) stated that the strong correlation (*r ≥* 0.970) between HRV and PRV measures in their study indicates that PRV could be a surrogate of HRV in stationary conditions. However, the authors do note that the PTT variability can cause slight differences between these two metrics, such that greater PTT variance would contribute to greater discrepancies. In support of this, Wong et al. found differences between both frequency- and time-domain HRV and PRV indices but also between the left and right PRV values, further highlighting that these methods are not interchangeable (15). It also seems plausible that positive publication bias may make it more likely for high correlations between PRV and HRV to be published as opposed to studies finding low or non-significant relationships (22–24).

Previous studies have provided preliminary insight on the issue of using HRV and PRV interchangeably, revealing some limitations. For example, the bulk of the literature has investigated HRV within healthy young adults, primarily males, which limits the generalizability to older (≥ 60 years) or younger (<18 years) individuals who may have altered PTT regulation. Additionally, the position of the PPG sensor differs across studies, including the finger, wrist or earlobe, as well as the ECG being conducted either centrally (traditional thorax setup) or peripherally (at forearm) (25). Further, each study has presented various HRV outcomes within the frequency or time domains, which contributes to inconsistent findings and reproducibility issues. With these research design inconsistencies and small sample sizes, more research is needed to understand the HRV-PRV relationship. When empirical data is scarce or of low quality but there is a known fundamental relationship between events (i.e., there is a causal relationship between electrical contraction of cardiac tissue and blood flow in the microvasculature), it is often helpful to use mathematical or statistical simulations to show how these events play out under various perturbations.

Thus, the purpose of this study was to use two large and publicly available datasets and Bayesian simulation principles to 1) determine if PPG-derived HRV (i.e., PRV) is equivalent to ECG-derived HRV and 2) if HRV and PRV are not equivalent, determine if the measurement characteristics of PRV are sufficiently viable for it to be a biomarker separate from HRV.

## Methods

### Raw ECG Data

Raw ECG data were downloaded from Physionet within the Autonomic Aging data set (26). The Waveform Database Software Package (WFDB) within MATLAB (MATLAB version: 9.13.0 (R2022b), Natick, Massachusetts: The MathWorks Inc.; 2022) was used to determine the correct channel and threshold for the ECG interbeat intervals (IBIs) for each participant. The histogram feature was used to ensure normal distribution of the R-R intervals with no outliers. If there were significant outliers or a non-normally distributed curve with the default settings (signal=1, threshold=1), a different signal (e.g., 2, 3) or threshold (e.g., 0.5, 2) was applied to obtain the best signal. The final output including all the IBIs was saved as a new text file for further analysis.

The annotated IBIs for the 1,084 patients were then exported at the millisecond resolution level (sampling rate = 1000 Hz) to the R programming language and analyzed using the RHRV package (27). Invalid IBIs were removed using an adaptive thresholding technique which has been previously described (28). A cleaned version of the QRS timings were then recovered for each participant for use in the planned simulation.

### Raw Pulse Transit Time Variability Data

Raw PTT data was extracted from open-source physiological data from the work of Mol et al. (29). In brief, supine rested PTT was collected in younger and older adults at the left wrist and index finger. The PTT was calculated as the time interval between the ECG derived R-wave and the peak of the first derivative PPG signal for both the wrist and finger. Individual PTT cycles were extracted for young adults (finger PTT n=30, wrist PTT n=28) and older adults (finger PTT n=19, wrist PTT n=14). Mean and standard deviation for 60s clips within each individual and measurement site were determined.

### Bayesian Simulation of PPG-based PRV Measures

The PTT variability was used to place a Bayesian prior on the arrival time of each QRS timing to convert it into the distal arrival of the PPG waveform at the location corresponding to the PTT variation. The magnitude of the PTT itself was not meaningful, just as the magnitude of absolute timing for the QRS is not meaningful, but rather the interval between these waveforms. Essentially, a Gaussian probability density function was centered at the QRS time and the standard deviation of that probability density function corresponded to the PTT variability from a participant in the Mol et al study (See Figure 2). After constructing these probability density functions for each QRS time, that participant’s ECG series was then sampled 10,000 times, creating 10,000 simulated ECG series for each patient for each PTT variance at each distal location (finger or wrist). The root mean square of successive differences (RMSSD) and the standard deviation of N-N intervals (SDNN) were then calculated for each simulated time series.

**Figure 2.**
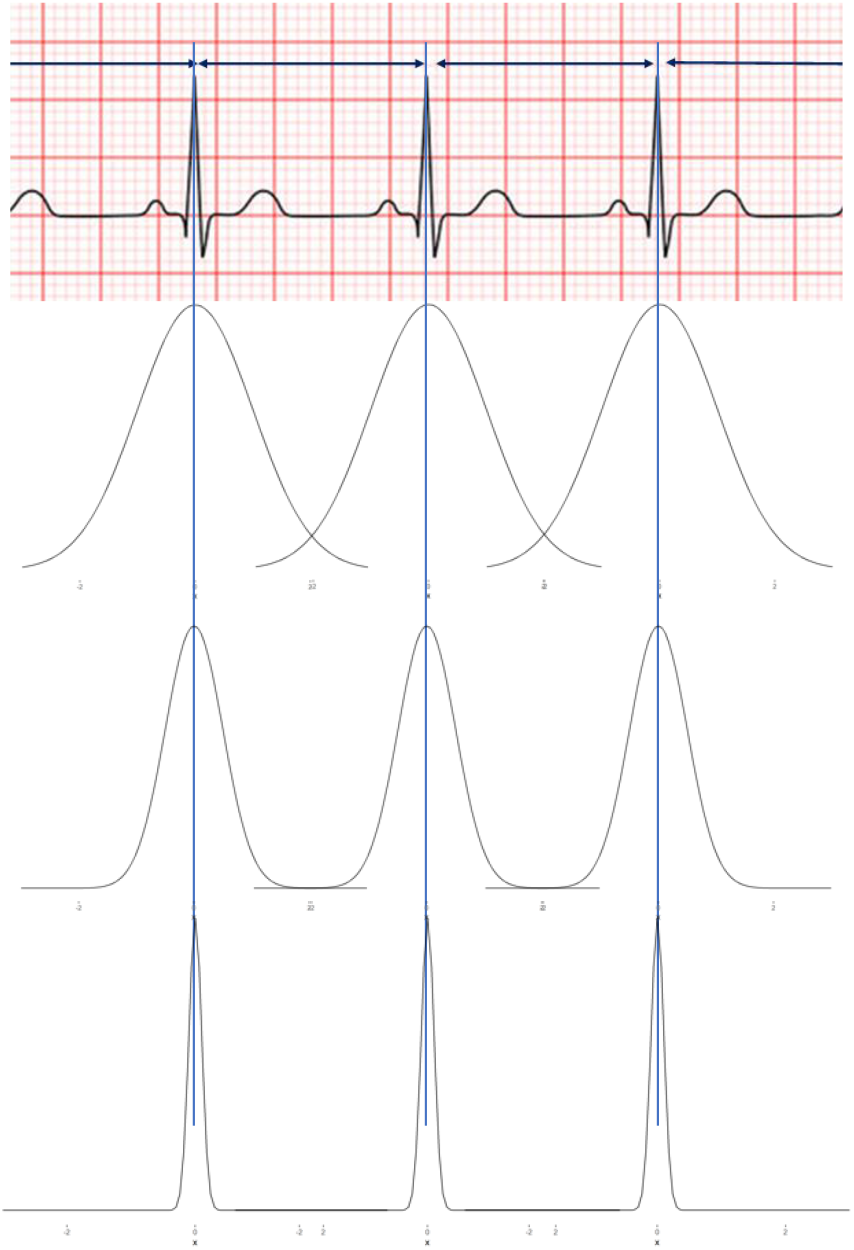
An example of the standard distribution of pulse rate values based on pulse transit time standard deviation (SD), with A representing a large SD and C representing a very small SD.

The primary goal of this work was to ascertain if HRV measured with ECG was equivalent to PRV measured by consumer wearables in the periphery. As such the Region of Practical Equivalence (ROPE) analysis was performed (30, 31). In the current work, the “real HRV” was a known quantity, so we simply needed to define what interval around the known quantity is acceptable to call HRV and PRV equivalent. To do this, we revisited previously obtained ECG measurements and determine how much variation one would expect to see between HRV metrics from ECG Lead II and Lead III recorded at the exact same time from the individuals within the Mol et al. study (29). This variation was slightly less than 0.1% between leads and so we empirically decided that it was generous to call any metric ±0.2% of the real HRV as “equivalent.” For each patient’s simulated PRV time series, we determined if it fell within the ROPE bounds. The percent of readings falling within ROPE can then be calculated and stratified by PTT variance to understand if PRV can ever be considered as equivalent to HRV and if it can, under what sort of peripheral cardiovascular constraints this is a valid assumption.

For each patient’s simulated time series at each PTT variance, the highest density interval (HDI) of the posterior was calculated for both RMSSD and SDNN. The HDI is an interval which encompasses 95% of the posterior distribution, is a measure of uncertainty and sometimes called the “credible interval” (32). This allows us to determine the most likely ranges of SDNN or RMSSD measures that would be obtained under ideal circumstances from a PPG sensor at a given location for a singular person. The width of the HDI is calculated for each patient within a given PTT variance and the median HDI for the PTT variance is reported. This allows us to know, for a standard ‘true HRV’, how much variance we would see in PRV if only peripheral cardiovascular variance is considered.

## Results

For the same PTT variance, SDNN had more HRV-PRV equivalent observations than RMSSD. For example, the lowest PTT variance observed was at the wrist and was 2.0 standard deviations, corresponding to 88.4% within ROPE for SDNN and 21.4% within ROPE for RMSSD. There is a clear relationship where lower PTT variance leads to higher ROPE, until ROPE becomes zero (Figure 3). For SDNN, ROPE becomes zero (zero equivalence between PRV and HRV) at 7.6 PTT standard deviations and at 5.5 PTT standard deviations for RMSSD.

**Figure 3.**
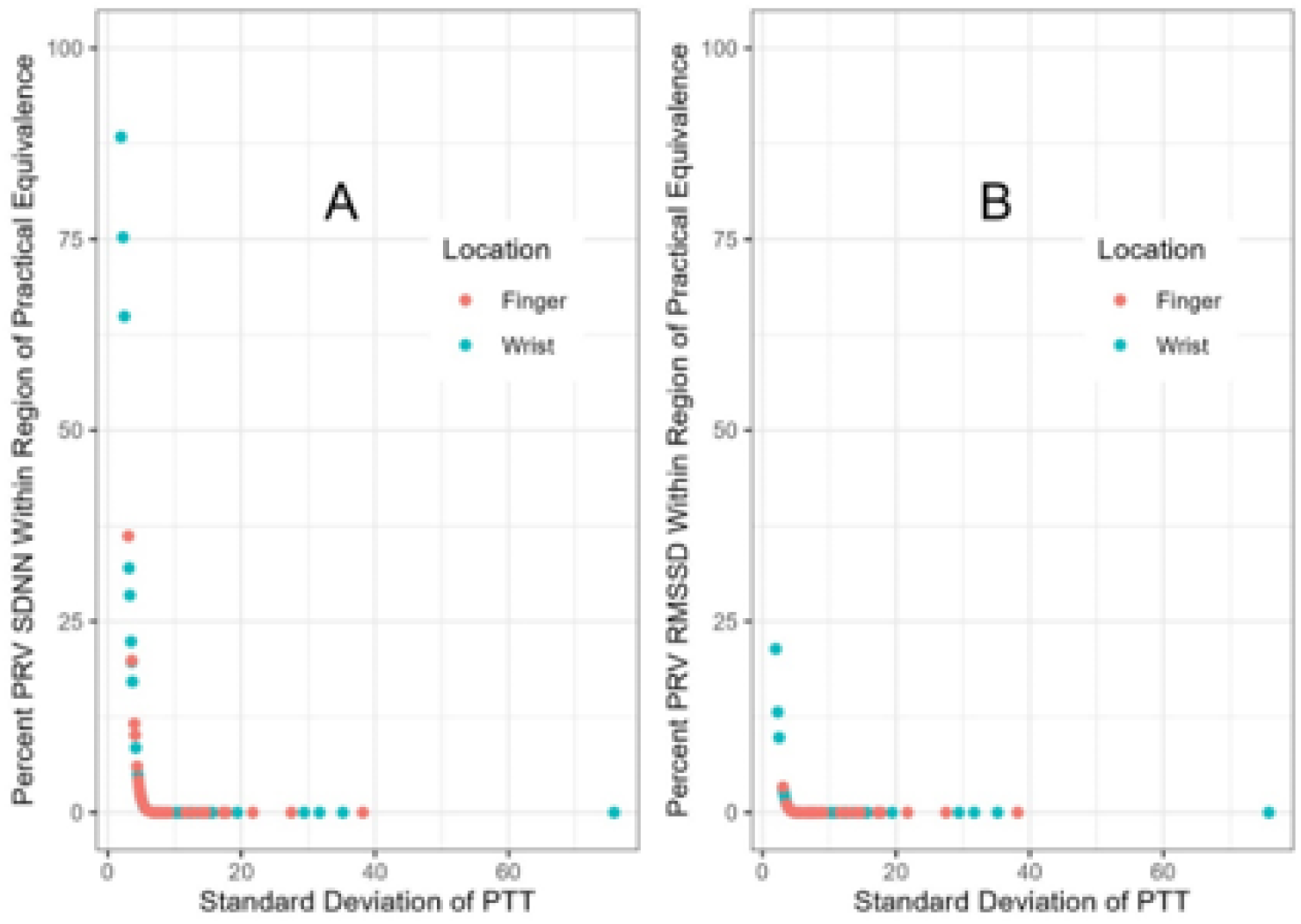
Results from the ROPE Analyses simulation. Plot A shows that PRV and HRV SDNN measures are only equivalent when PTT variance is low. Plot B shows that the PRV and HRV RMSSD metric are generally not equivalent at the 0.2% level.

This demonstrates that SDNN has a very narrow band of PTT variance where PRV and HRV are broadly equivalent and RMSSD has nearly no likely real-world equivalence between PRV and HRV.

For both SDNN and RMSSD, the HDI interval width increases with increasing PTT variance. However, the dynamics of this change appear different between SDNN (Figure 4A) and RMSSD (Figure 4B). The rate of HDI width, a measure of uncertainty, increases at a far higher rate for RMSSD than it does for SDNN. For example, the HDI width of SDNN does not reach 1 unit until PTT variance is 10 standard deviations, whereas RMSSD HDI width is 1 unit at 3.5 standard deviations.

**Figure 4.**
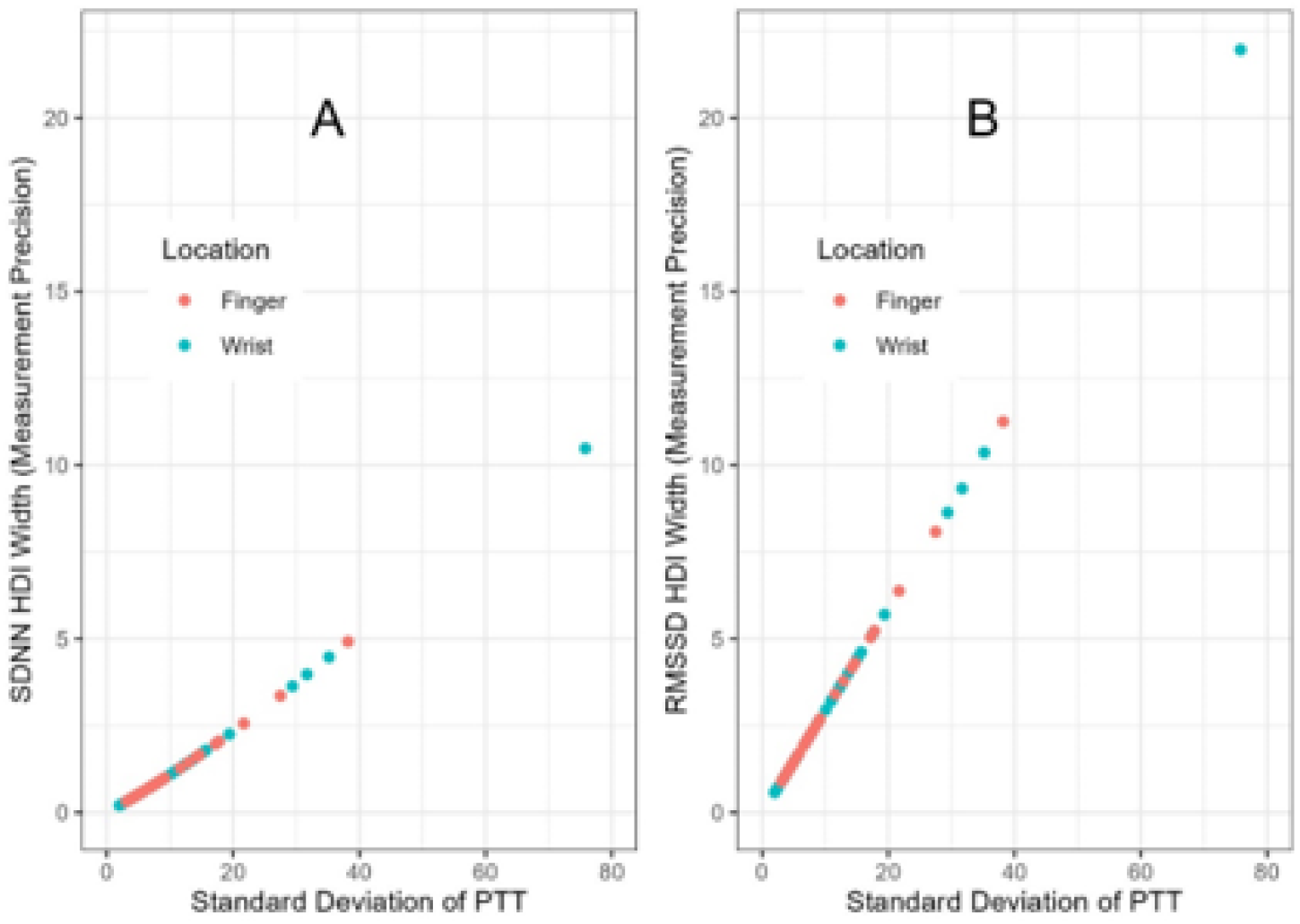
Results from the HDI analyses showing that both SDNN (Plot A) and RMSSD (Plot B) have increasing measurement error with increases in PTT variance. The slope of the increase is more profound for RMSSD than SDNN.

## Discussion

Our study found that greater PTT variance not only decreased the equivalence of PRV to HRV, but also increased the measurement error of PRV itself. More specifically, PTT variance should be less than two standard deviations for SDNN to be deemed equivalent (i.e., within 80% ROPE); however, HRV-PRV RMSSD showed no equivalence with any amount of PTT variance. Ultimately, this indicates that ECG-derived HRV and PPG-derived PRV should not be used interchangeably when defining “HRV” as a general metric. When considering PRV as a unique biometric measure, our findings reveal that SDNN has more favorable measurement properties than RMSSD, though both exhibit a non-uniform measurement error.

Recently, both peripherally located PPG and centrally located ECG have been utilized to measure “HRV”. A review by Mejia-Mejia et al. (19) indicated that even if pulse rate and heart rate can be used interchangeably, PRV and HRV refer to the variability around the mean and may not be similar estimates of each other. As outlined by Yuda and colleagues (16), there are several physiological steps between the electrical wave (QRS complex) at the heart and the mechanical pulse wave at the periphery, which may contribute to a discrepancy between the two. One specific factor that the authors mentioned is defined as PTT, with the speed of the pulse wave influenced by arterial radius, wall thickness and elasticity, and blood density (16). Further, these factors may be subject to change within various populations and situations (e.g., age, sex, activity, posture), thereby creating disconnect between heart rate and pulse rate (19, 20, 33). In support of this, the present study confirms that PRV and HRV may not be equivalent with greater PTT variance.

Previous studies have identified particular influences on PTT, which contribute to the dissociation of PRV from HRV. For example, aging is associated with decreased vascular elasticity, which may reduce PTT variance and enhance the relationship between HRV and PRV (21). Healthy and active (versus sedentary) individuals can exhibit improved vascular elasticity (34, 35), which may indicate that PRV is less reliable in these individuals. Furthermore, respiration (spontaneous vs. paced) (36), position (supine vs. head-up) (21), and exercise (37, 38) can also influence the relationship between HRV and PRV through alterations in PTT. Previous work has also found interindividual variability within the pre-ejection period, defined as the time between the Q-wave and the actual blood ejection from the heart, that can occur in response to stress conditions or the location of the peripheral sensor (39, 40).

This study chose to examine SDNN and RMSSD, as these are the most common time-domain variables assessed with wearable health monitoring devices (e.g., Oura, Apple Watch). Previous research has shown that RMSSD evaluates short-term dynamics, while SDNN may reflect more long-term properties (2). Our results indicate that SDNN may be a more stable and less error-prone measure of PRV than RMSSD, with little equivalence between ECG-derived and PPG-derived RMSSD regardless of PTT variance. One factor may be that RMSSD is more sensitive to artifacts than other HRV metrics, as suggested by Bourdillon and colleagues (41). Wearable technology or research studies utilizing PPG-derived variability metrics may consider using variables less influenced by short-term changes or artifact, such as SDNN. Further work is needed to explore this relationship within frequency-domain variables.

The outcome of our simulation is valid because the two causal events, heart electrical waveform (QRS complex) and peripheral pulse wave arrival, can be directly linked and simulated with physiologically-based data (PTT variance). Regarding the statistical analyses, it is important to note that a ROPE value of ±0.2% was chosen based on previous data. We would expect up to approximately ±0.1% discrepancy in HRV within the same individual during the same time window, as explained above, and equivalence should be characterized by a narrow range set *a priori* and not adjusted to ‘fit the data’ *post hoc*. Additionally, a measure must be considered consistent to be clinically meaningful or utilized to detect significant changes. Our HDI analyses held the “real HRV” constant and varied only the PTT variance, showing that both SDNN and RMSSD have systematically varying measurement error, but that this error is most profound in RMSSD. Certainly, there are other potential physiological processes which affect the measurement of PRV, but until the impacts of those processes can be effectively modeled, RMSSD PRV should be avoided and SDNN PRV should be viewed skeptically if PTT variance cannot be assumed to be constant. Although PPG-derived PRV metrics are widely used and may offer practical utility, our findings emphasize the need for clarification on terminology within HRV research to limit the interchangeability of HRV and PRV. Further, it should not be assumed that the inferences made by previous HRV research will translate to PRV findings.

## Data Availability

All data used in this study was publicly available at the following locations:
Pulse Transit Time from Mol et al: http://hdl.handle.net/11633/aacthxia
Autonomic Aging Dataset: https://physionet.org/content/autonomic-aging-cardiovascular/1.0.0/

http://hdl.handle.net/11633/aacthxia

